# Development and validation of the Elecsys Anti-SARS-CoV-2 immunoassay as a highly specific tool for determining past exposure to SARS-CoV-2

**DOI:** 10.1101/2020.06.16.20132803

**Authors:** Peter Muench, Simon Jochum, Verena Wenderoth, Beatus Ofenloch-Haehnle, Michael Hombach, Matthias Strobl, Henrik Sadlowski, Christopher Sachse, Alexander Riedel

## Abstract

**Background:** The Elecsys^®^ Anti-SARS-CoV-2 immunoassay (Roche Diagnostics) was developed to provide an accurate and reliable method for the detection of antibodies to severe acute respiratory syndrome coronavirus 2 (SARS-CoV-2). We evaluated the sensitivity, specificity, and cross-reactivity of the Elecsys Anti-SARS-CoV-2 immunoassay.

**Methods:** The performance of the Elecsys Anti-SARS-CoV-2 immunoassay was assessed at Roche Diagnostics (Penzberg, Germany). Sensitivity was evaluated using anonymised residual frozen samples from patients who had previously tested positive for SARS-CoV-2 infection by polymerase chain reaction (PCR); one or more consecutive samples were collected from patients at various timepoints after PCR confirmation. Specificity was evaluated using anonymised unselected residual frozen samples from routine diagnostic testing or from blood donors; all samples were collected before December 2019 and thus deemed negative for SARS-CoV-2-specific antibodies. Cross-reactivity was evaluated using anonymised frozen samples containing a wide range of potentially cross-reacting analytes, which were purchased from commercial vendors. For sensitivity and specificity, point estimates and 95% confidence intervals (CIs) were calculated.

**Results:** Sensitivity of the Elecsys Anti-SARS-CoV-2 immunoassay in 496 samples from 102 patients with prior PCR-confirmed SARS-CoV-2 infection was 99.5% (95% CI 97.0–100.0) at ≥14 days after PCR confirmation. Overall specificity in 10,453 samples from routine diagnostic testing (n = 6305) and blood donors (n = 4148) was 99.80% (95% CI 99.69–99.88). Only 4/752 samples containing potential cross-reacting analytes were reactive with the Elecsys Anti-SARS-CoV-2 immunoassay, resulting in an overall specificity in this cohort of 99.5% (95% CI 98.6–99.9).

**Conclusion:** The Elecsys Anti-SARS-CoV-2 immunoassay demonstrated a sensitivity of 99.5% at ≥14 days after PCR confirmation and a very high specificity of 99.80%. Our findings support the use of the Elecsys Anti-SARS-CoV-2 immunoassay as a tool for the identification of past SARS-CoV-2 infection, including in populations with a low disease prevalence.

**Required information for submission system:** *Ethical guidelines:* The study was conducted in accordance with applicable regulations, including relevant European Union directives and regulations, and the principles of the Declaration of Helsinki. All samples, excluding the specimens that were provided by commercial sample vendors, were transferred to Roche following anonymisation. For studies with anonymised leftover specimens, no ethics committee vote is required. A statement was obtained from the Ethics Committee of the Landesä rztekammer Bayern confirming that there are no objections against the transfer and the coherent use of the anonymised leftover samples.

*Research reporting guidelines:* Please see separate STARD checklist

*Data availability statement:* Qualified researchers may request access to individual patient level data through the clinical study data request platform (https://vivli.org/). Further details on Roche’s criteria for eligible studies are available here: https://vivli.org/members/ourmembers/. For further details on Roche’s Global Policy on the Sharing of Clinical Information and how to request access to related clinical study documents, see here: https://www.roche.com/research_and_development/who_we_are_how_we_work/clinical_trials/our_commitment_to_data_sharing.htm.

## Introduction

In December 2019, reports emerged of patients presenting with pneumonia of unknown aetiology in Wuhan, China.^1,2^ This disease was subsequently shown to be caused by a novel coronavirus, severe acute respiratory syndrome coronavirus 2 (SARS-CoV-2), and designated coronavirus disease 2019 (COVID-19).^3–5^ Since then, the COVID-19 outbreak has rapidly developed into a pandemic, which has infected millions of people and posed critical challenges for governments and healthcare systems around the world.^6^

SARS-CoV-2 is an enveloped, single-stranded RNA virus of the *Coronaviridae* family. All coronaviruses share similarities in the organisation and expression of their genome, which encodes 16 non-structural proteins and four structural proteins: spike, envelope, membrane, and nucleocapsid antigens.^5,7–9^ Evidence to date suggests that SARS-CoV-2 is primarily transmitted between people through respiratory droplets and contact routes, although indirect transmission via contaminated surfaces is also possible.^10–12^ Infected individuals may exhibit a variety of symptoms, including fever, cough, and breathlessness, and disease severity can range from asymptomatic/mild cases to severe disease and death.^13,14^

There is an urgent unmet clinical need to more effectively determine SARS-CoV-2 seroprevalence in the general population to improve our understanding of virus circulation dynamics, gain a more accurate estimate of the mortality rate from COVID-19, and identify individuals at risk of infection. Serological assays for SARS-CoV-2 have been suggested as a potential tool to help identify the extent of virus exposure in a given population and thereby indirectly inform on the appropriate application, enforcement, or relaxation of containment measures.^15–18^ Serological assays may also help elucidate a potential correlate for subsequent immunity following infection.^15,16^

The timing of seroconversion is crucial for determining optimum timepoints for sample collection for serological testing.^19^ Although the picture is rapidly developing and robust serology data are not yet available, antibody kinetics to SARS-CoV-2 have begun to be described. Based on current evidence, immunoglobulin M (IgM) antibodies are detectable within 5 days after symptom onset and immunoglobulin G (IgG) antibodies within 5–7 days.^20–22^ There is a paucity of data on immunoglobulin A (IgA), but it appears to be observable approximately 3–6 days after symptom onset.^15,21^ Depending on the applied method, seroconversion is observed after a median of 10–13 days after symptom onset for IgM and 12–14 days for IgG; maximum seroconversion occurs at 2–3 weeks for IgM, 3–6 weeks for IgG, and 2 weeks for total antibodies.^20,22–26^ Levels and chronological order of IgM and IgG antibody appearance are highly variable, supporting detection of both antibodies simultaneously.^17,22,23,25^

The Elecsys^®^ Anti-SARS-CoV-2 immunoassay (Roche Diagnostics International Ltd, Rotkreuz, Switzerland) was developed to provide an accurate and reliable method for the detection of antibodies to SARS-CoV-2, in order to facilitate population screening with high specificity and the identification of past infection status as a potential correlate for subsequent immunity. We aimed to evaluate the sensitivity, specificity, and cross-reactivity of the Elecsys Anti-SARS-CoV-2 immunoassay.

## Materials and Methods

### Study design

The performance of the Elecsys Anti-SARS-CoV-2 immunoassay was prospectively evaluated at Roche Diagnostics (Penzberg, Germany). Sensitivity and specificity analyses were conducted using anonymised residual frozen samples from routine diagnostic testing or from blood donors, which were obtained from diagnostic laboratories in Germany and a blood product provider in the United States, respectively. Cross-reactivity analyses were conducted using anonymised frozen samples containing potentially cross-reacting factors, which were purchased from commercial vendors.

The study was conducted in accordance with applicable regulations, including relevant European Union directives and regulations, and the principles of the Declaration of Helsinki. All samples, excluding the specimens that were provided by commercial sample vendors, were transferred to Roche following anonymisation. For studies with anonymised leftover specimens, no ethics committee vote is required. A statement was obtained from the Ethics Committee of the Landesärztekammer Bayern confirming that there are no objections against the transfer and the coherent use of the anonymised leftover samples.

### Assay

The Elecsys Anti-SARS-CoV-2 electrochemiluminescence immunoassay is intended for use on cobas e analysers (Roche Diagnostics International Ltd, Rotkreuz, Switzerland) for the in-vitro qualitative detection of antibodies (including IgG) to SARS-CoV-2 in human serum and plasma. The immunoassay utilises a double-antigen sandwich test principle and a recombinant protein representing the nucleocapsid antigen for the determination of antibodies to SARS-CoV-2.

For the present study, the Elecsys Anti-SARS-CoV-2 immunoassay was performed according to manufacturer instructions and assay results were interpreted as follows: cut-off index <1.0, non-reactive/negative for anti-SARS-CoV-2 antibodies; cut-off index ≥1.0, reactive/positive for anti-SARS-CoV-2 antibodies.

### Sensitivity

Sensitivity of the Elecsys Anti-SARS-CoV-2 immunoassay was evaluated using residual samples from patients who had previously tested positive for SARS-CoV-2 infection by polymerase chain reaction (PCR). One or more consecutive samples were collected from patients at various timepoints after PCR confirmation. Samples derived from all patients with prior PCR-confirmed SARS-CoV-2 infection were used; no additional sample selection was made except for the availability of a sufficient amount of residual serum or plasma. Assay sensitivity at different timepoints after PCR confirmation was calculated as the percentage of samples that tested positive with the Elecsys Anti-SARS-CoV-2 immunoassay relative to the total number of PCR-confirmed positive samples.

### Specificity

Specificity of the Elecsys Anti-SARS-CoV-2 immunoassay was evaluated using unselected residual samples from routine diagnostic testing or from blood donors. All samples were collected before December 2019 (i.e. before the first description of an infection with SARS-CoV-2) and were thus deemed negative for SARS-CoV-2-specific antibodies. Assay specificity was calculated as the percentage of true SARS-CoV-2 antibody-negative samples that tested negative with the Elecsys Anti-SARS-CoV-2 immunoassay.

### Cross-reactivity

Cross-reactivity of the Elecsys Anti-SARS-CoV-2 immunoassay was evaluated using samples that had previously been characterised as positive for a wide range of different indications, including a common cold panel and samples from patients with autoimmune conditions and other diseases that are associated with a higher prevalence of autoantibodies and immune dysfunction, which may increase the risk of interference with serological testing. Assay specificity was calculated for each potential cross-reactive sample type and for the total cohort as the percentage of samples tested that were reactive with the Elecsys Anti-SARS-CoV-2 immunoassay.

### Statistical analyses

Sample size estimation for the specificity analyses was calculated using the binomial exact test.^27^ The following sample sizes for a seronegative cohort would be required to obtain alpha = 5% and power = 80%: p0 = 0.99, n = 368; p0 = 0.995, n = 736; p0 = 0.996, n = 921; p0 = 0.997, n = 1228; p0 = 0.998, n = 1843; p0 = 0.999, n = 3688; p0 = 0.9995, n = 7376. A sample size of n >10,000 would ensure the analyses were sufficiently powered to provide a reliable estimate of assay specificity. For the cross-reactivity analyses, the aim was to evaluate n ≥10 samples per cross-reacting factor.

Determinations were performed as single measurements. For sensitivity and specificity, point estimates and 95% confidence intervals (CIs) were calculated.

## Results

### Sensitivity

A total of 496 samples from 102 patients with prior PCR-confirmed SARS-CoV-2 infection were included in the sensitivity analyses. Sensitivity increased with time after PCR confirmation: 60.2% (95% CI 52.3–67.8) at 0–6 days; 85.3% (95% CI 78.6–90.6) at 7–13 days; and 99.5% (97.0–100.0) at ≥14 days (**Table 1**; **Figure 1**).

**Table 1.** Summary of sensitivity results for the Elecsys Anti-SARS-CoV-2 immunoassay in patients with prior PCR-confirmed SARS-CoV-2 infection.

| Days after PCR confirmation | Samples tested, n | Reactive samples, n | Non-reactive samples, n | Sensitivity, % (95% confidence interval) |
| --- | --- | --- | --- | --- |
| 0–6 | 161 | 97 | 64 | 60.2 (52.3–67.8) |
| 7–13 | 150 | 128 | 22 | 85.3 (78.6–90.6) |
| ≥14 | 185 | 184 | 1* | 99.5 (97.0–100.0) |

\*One patient was non-reactive at Day 14 (cut-off index 0.7) but reactive at Day 16 (cut-off index 4.5).
PCR, polymerase chain reaction; SARS-CoV-2, severe acute respiratory syndrome coronavirus 2.

**Figure 1.**
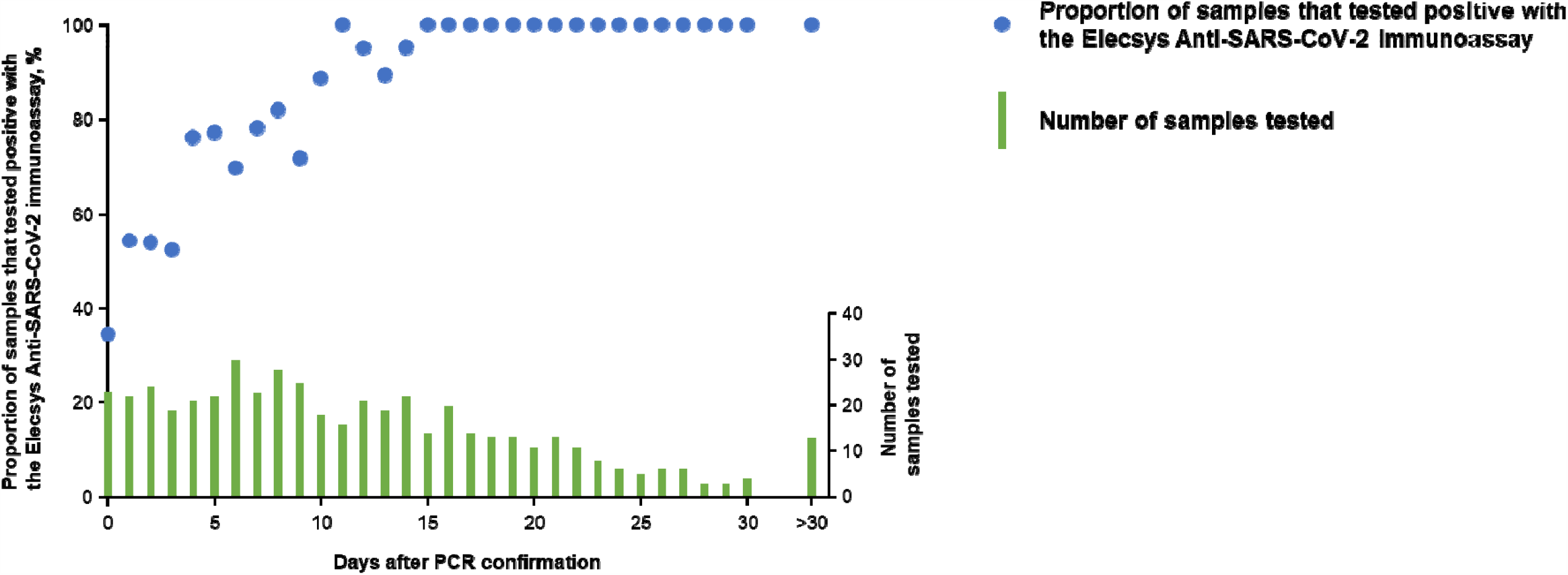
Sensitivity of the Elecsys Anti-SARS-CoV-2 immunoassay in patients with prior PCR-confirmed SARS-CoV-2 infection. PCR, polymerase chain reaction; SARS-CoV-2, severe acute respiratory syndrome coronavirus 2.

Elecsys Anti-SARS-CoV-2 immunoassay results for 26 consecutive samples from five patients following recovery from PCR-confirmed SARS-CoV-2 infection are shown in **Figure 2**.

**Figure 2.**
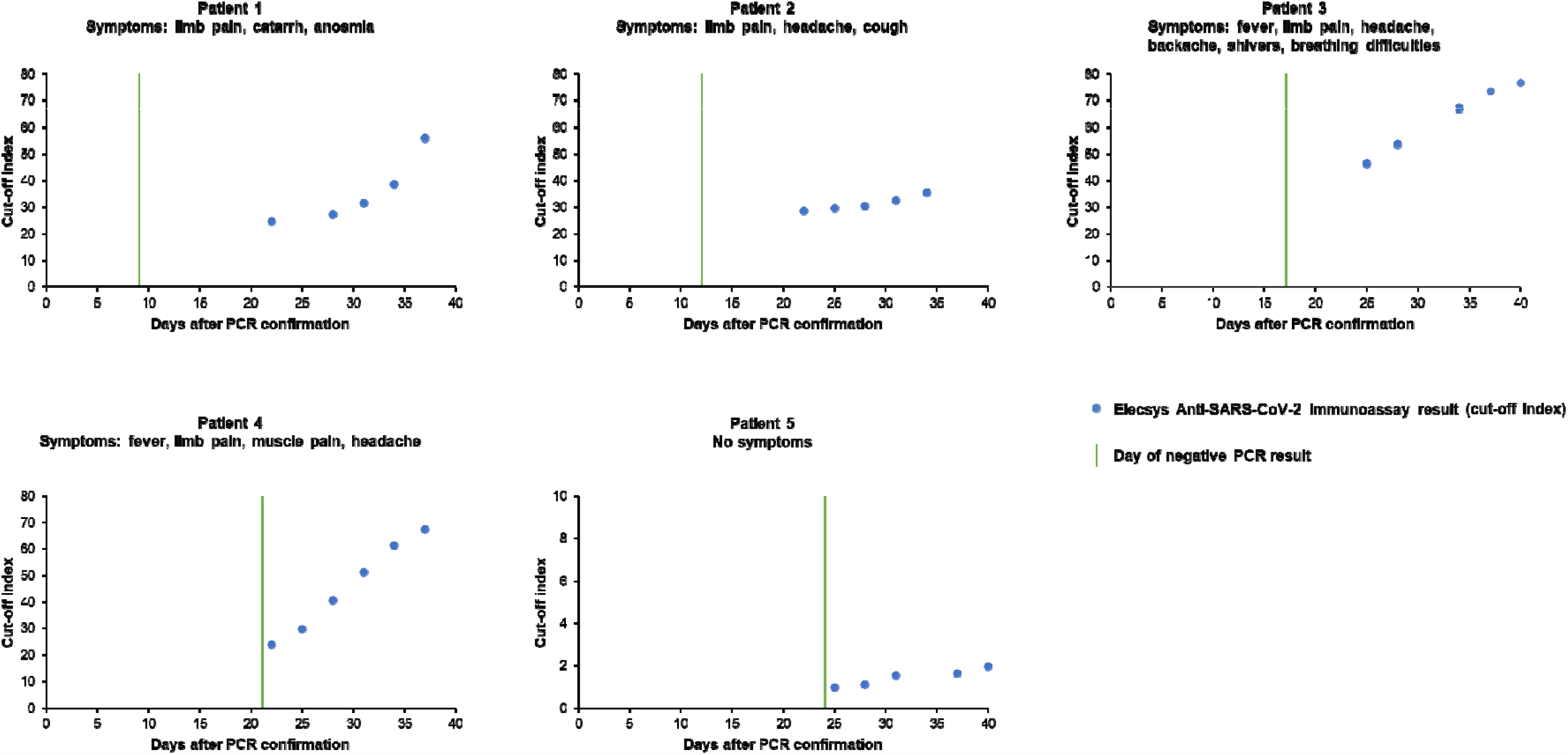
Elecsys Anti-SARS-CoV-2 immunoassay results in five patients following recovery from PCR-confirmed SARS-CoV-2 infection. Day 0 represents day of initial positive PCR result. PCR, polymerase chain reaction; SARS-CoV-2, severe acute respiratory syndrome coronavirus 2.

### Specificity

A total of 10,453 samples were included in the specificity analyses: 6305 samples from routine diagnostic testing and 4148 samples from blood donors. The overall specificity for the entire sample cohort was 99.80% (95% CI 99.69–99.88); 99.81% (95% CI 99.67–99.90) in routine diagnostic samples and 99.78% (95% CI 99.59–99.90) in blood donor samples (**Table 2**).

**Table 2.** Summary of specificity results for the Elecsys Anti-SARS-CoV-2 immunoassay in residual samples from routine diagnostic testing and blood donors.

| Sample cohort | Samples tested, n | Reactive samples, n | Non-reactive samples, n | Specificity, % (95% confidence interval) |
| --- | --- | --- | --- | --- |
| Routine diagnostic samples | 6305 | 12 | 6293 | 99.81 (99.67–99.90) |
| Blood donor samples | 4148 | 9 | 4139 | 99.78 (99.59–99.90) |
| **Total** | **10,453** | **21** | **10,432** | **99.80 (99.69–99.88)** |

### Cross-reactivity

Out of 752 samples with potential cross-reactivity, four samples were reactive with the Elecsys Anti-SARS-CoV-2 immunoassay: 1/85 (1.2%) acute cytomegalovirus infection samples; 2/105 (1.9%) acute Epstein-Barr virus infection samples; and 1/10 (10.0%) systemic lupus erythematosus samples (**Table 3**). The overall specificity in this cohort was 99.5% (95% CI 98.6–99.9; **Table 3**).

**Table 3.** Summary of cross-reactivity results for the Elecsys Anti-SARS-CoV-2 immunoassay.

| Potential cross-reactive sample type | Samples tested, n | Reactive samples, n | Specificity, % |
| --- | --- | --- | --- |
| Common cold panel | 40 | 0 | 100.0 |
| Cytomegalovirus – acute infection (IgM/IgG positive) | 85 | 1 | 98.8 |
| Epstein-Barr virus – acute infection (IgM/IgG positive) | 105 | 2 | 98.1 |
| *Borrelia* | 6 | 0 | 100.0 |
| *Chlamydia pneumoniae* | 8 | 0 | 100.0 |
| *Escherichia coli* (anti-*E. coli* reactive) | 10 | 0 | 100.0 |
| Gonorrhoea | 5 | 0 | 100.0 |
| Hepatitis A virus – acute infection (IgM positive) | 10 | 0 | 100.0 |
| Hepatitis A virus – late infection (IgG positive) | 15 | 0 | 100.0 |
| Hepatitis A virus vaccinees | 15 | 0 | 100.0 |
| Hepatitis B virus – early acute infection (HBsAg/HBeAg positive) | 12 | 0 | 100.0 |
| Hepatitis B virus – acute infection (anti-HBs positive) | 7 | 0 | 100.0 |
| Hepatitis B virus – acute infection (anti-HBc IgM positive) | 8 | 0 | 100.0 |
| Hepatitis B virus – chronic infection | 12 | 0 | 100.0 |
| Hepatitis B virus vaccinees | 15 | 0 | 100.0 |
| Hepatitis C virus – acute infection (IgM positive) | 6 | 0 | 100.0 |
| Hepatitis C virus (IgG positive) | 60 | 0 | 100.0 |
| Hepatitis E virus | 12 | 0 | 100.0 |
| Human immunodeficiency virus | 10 | 0 | 100.0 |
| Herpes simplex virus – acute infection (IgM positive) | 24 | 0 | 100.0 |
| Human T-lymphotropic virus | 6 | 0 | 100.0 |
| Influenza vaccinees | 25 | 0 | 100.0 |
| Listeria | 6 | 0 | 100.0 |
| Measles | 10 | 0 | 100.0 |
| Mumps | 14 | 0 | 100.0 |
| Parvovirus B19 | 30 | 0 | 100.0 |
| *Plasmodium falciparum* (malaria) | 8 | 0 | 100.0 |
| Rubella – acute infection (IgM/IgG positive) | 12 | 0 | 100.0 |
| *Toxoplasma gondii* (IgM/IgG positive) | 8 | 0 | 100.0 |
| *Treponema pallidum* (syphilis) | 62 | 0 | 100.0 |
| Varicella zoster virus | 30 | 0 | 100.0 |
| Anti-mitochondrial antibodies | 30 | 0 | 100.0 |
| Anti-nuclear antibodies | 26 | 0 | 100.0 |
| Systemic lupus erythematosus | 10 | 1 | 90.0 |
| Rheumatoid arthritis | 10 | 0 | 100.0 |
| **Total** | **752** | **4** | **99.5 (95% CI 98.6–99.9)** |

Anti-HBc, antibodies to hepatitis B core antigen; Anti-HBs, antibodies to hepatitis B surface antigen; CI, confidence interval; IgG, immunoglobulin G; IgM, immunoglobulin M; HBeAg, hepatitis B envelope antigen; HBsAg, hepatitis B surface antigen.

## Discussion

The COVID-19 pandemic has resulted in significant mortality and morbidity, and created major challenges for governments and healthcare systems. Due to the novelty of the causative agent, SARS-CoV-2, and the rapidity with which it has spread, there is an urgent need for serological assays, which could be used to determine the seroprevalence in a given population and help identify a potential correlate for immunity secondary to exposure.^15–18^ This urgency and the lack of existing available tests prompted the US Food and Drug Administration to allow manufacturers of serological assays for SARS-CoV-2 to bypass the normal approval process, resulting in an inundation of tests being released into the market, many of which have not been sufficiently reviewed by regulatory authorities nor their performance reported in the peer-reviewed literature.^16^ However, it is vital that any new test is adequately evaluated and validated to demonstrate that it is reliable and accurate for its intended purpose.

To enable population screening to determine the level of exposure and identify individuals who may be immune following infection, an ideal serological test for SARS-CoV-2 should meet several key requirements: 1) a high general specificity with a small CI; 2) no cross-reactivity with other endemic coronaviruses; 3) preference for the detection of mature antibodies to provide the potential for correlation with neutralising activity; and 4) a high sample throughput to meet the huge demand for testing. The first three requirements can be achieved through the application of appropriate assay format and antigen selection, while the last requirement relies on a high-throughput platform and upscaling of laboratories. Although a high assay sensitivity is desirable, it is of secondary importance and should not be pursued at the expense of specificity for past infection, as a serological assay is less likely to be used for the diagnosis of active infection.

The Elecsys Anti-SARS-CoV-2 immunoassay was specifically designed to meet these requirements. It utilises a double-antigen sandwich test principle for the detection of high-affinity (i.e. late-onset/mature) antibodies to SARS-CoV-2. In the present study, the Elecsys Anti-SARS-CoV-2 immunoassay demonstrated an overall specificity of 99.80% (95% CI 99.69–99.88) in 10,453 residual samples from routine testing and blood donors. The sensitivity of the Elecsys Anti-SARS-CoV-2 immunoassay in samples from patients with prior PCR-confirmed SARS-CoV-2 infection increased with time after PCR confirmation, reaching 99.5% (95% CI 97.0–100.0) at ≥14 days. The performance of the Elecsys Anti-SARS-CoV-2 immunoassay is comparable or better than that observed for other serological SARS-CoV-2 assays (specificity, 94.8–99.9%; sensitivity at ≥14 days post-PCR confirmation, 75.0–100.0%).^28,29^

The main risk with the use of serological SARS-CoV-2 assays for population screening is the possibility of false-positive results, which could lead to the erroneous assumption of past infection and subsequent putative immunity, and put the individual at risk of acquiring or transmitting infection.^16^ A very high specificity is crucial to reduce the rate of false-positive results, particularly in populations with low seroprevalence where small differences in assay specificity can result in substantial differences in the positive predictive value (PPV).^15,16^ For example, if the seroprevalence of SARS-CoV-2 in a given population was 10%, a serological test with a sensitivity of 83.1% and a specificity of 98.3% would have a PPV of 84.2%; however, if the seroprevalence is 1%, the respective PPV would drop to 32.6%.^30^ Understanding this relationship between seroprevalence, specificity, and PPV is crucial for population screening using serological testing, and it has been suggested that assay specificity should be >99% (with a CI of 99.0–99.9) to ensure a sufficient PPV in populations with low seroprevalence.^16^ Based on a sensitivity of 99.5% and a specificity of 99.8%, the PPV of the Elecsys Anti-SARS-CoV-2 immunoassay in populations of 1%, 5%, 10%, and 20% seroprevalence would be 83.4%, 96.3%, 98.2%, and 99.2%, respectively. A potential cause of false positives is cross-reactivity with other analytes. In the present study, only 4/752 samples containing potential cross-reacting analytes showed reactivity with the Elecsys Anti-SARS-CoV-2 immunoassay, resulting in an overall specificity of 99.5%.

Based on our understanding of other respiratory viruses, it has been suggested that the presence of anti-SARS-CoV-2 antibodies will provide some immunity, although it is unclear how long any protection would last for after initial infection.^31^ Assuming anti-SARS-CoV-2 antibodies do offer some immunity from further infection, donated plasma from patients who have recovered from COVID-19 may represent a potential therapy by conferring immunity to the recipient.^31^ In this context, serological assays could provide another important role in the identification of potential convalescent plasma donors, particularly individuals with very high anti-SARS-CoV-2 antibody titres.^16^ Recent studies suggest that both cell-mediated and humoral immune responses are likely to play a protective role in SARS-CoV-2 infection, and the spike and nucleocapsid antigens in particular have been shown to be highly immunogenic and abundantly expressed during infection.^7–9,32^ Antibodies targeting these proteins are formed as early as 9 days after symptom onset and have demonstrated a strong neutralising response, suggesting that seroconversion may lead to immunity for a limited time after infection.^15,17,33^ However, further research is needed to demonstrate the correlation between the presence of anti-SARS-CoV-2 antibodies and neutralisation, and ultimately the correlation with clinical immunity. Additional work is also required to establish whether there is a correlation between anti-SARS-CoV-2 antibody titres and disease severity and/or prognosis.^34^

A major strength of this study is the large number of seronegative samples (n = 10,453) used to determine the specificity of the Elecsys Anti-SARS-CoV-2 immunoassay, which ensures the study is robustly powered and the reliability of the specificity point estimates is very high (as indicated by the small CIs).^16,35^ To our knowledge, this is one of the largest sample sizes used to date to evaluate the specificity of a serological assay for SARS-CoV-2. Another study strength is the inclusion of a high number of confounder samples in the cross-reactivity cohort, which comprised a total of 752 samples across 35 different indications. A limitation is that this was a single-centre study and our results should be confirmed by additional assessments at other study sites. Further clinical data on the samples were not available due to data regulations, and thus it was not possible to analyse specific sub-cohorts according to age, disease severity, onset of symptoms, etc. We are confident that our results regarding the sensitivity of the Elecsys Anti-SARS-CoV-2 immunoassay and the time course of antibody response are of general value. However, the findings should be interpreted with some caution. Although the samples used in this study were not selected on the basis of patient criteria, the majority of samples were drawn from hospitalised patients and therefore probably represent more severe cases of COVID-19. Furthermore, the availability of samples depended on the need of consequent routine clinical chemistry diagnosis after PCR-confirmation of SARS-CoV-2 infection. Again, it can be assumed that extensive subsequent diagnosis was predominantly performed in cases with more severe disease. The validity of our findings in ambulatory settings or in patients with asymptomatic/mildly symptomatic SARS-CoV-2 infection is yet to be shown and requires further study.

## Conclusion

The Elecsys Anti-SARS-CoV-2 immunoassay demonstrated a very high specificity of 99.80% and a sensitivity of 99.5% for past infection in patients at ≥14 days after PCR confirmation, supporting its use as a potential tool for the identification of past exposure to SARS-CoV-2 infection. High assay specificity and sensitivity are crucial to ensure a high PPV for population screening, particularly in settings with low disease prevalence.

## Data Availability

Qualified researchers may request access to individual patient level data through the clinical study data request platform (https://vivli.org/). Further details on Roche's criteria for eligible studies are available here: https://vivli.org/members/ourmembers/. For further details on Roche's Global Policy on the Sharing of Clinical Information and how to request access to related clinical study documents, see here: https://www.roche.com/research_and_development/who_we_are_how_we_work/clinical_trials/our_commitment_to_data_sharing.htm.

## Acknowledgements

The authors thank the following individuals for their contributions to the study: Melanie Ermlich (Roche Diagnostics; support with sample acquisition and literature review); Zoey Germuskova (Roche Diagnostics; support with sample acquisition, defining the research question, and literature review); Manuela Haslinger (Roche Diagnostics; support with conducting specificity and cross-reactivity experiments); Victor Jeger (Roche Diagnostics; critical input); Florina Langen (Roche Diagnostics; support with sample acquisition); Peter Mackeben (Roche Diagnostics; support with conducting specificity and cross-reactivity experiments); Sigrid Reichhuber (Roche Diagnostics; support in sample acquisition); Tanja Schneider (Roche Diagnostics; support in sample acquisition); Maria von Holtey (Roche Diagnostics; support in sample acquisition, defining the research question, and literature review); Mirko Walter (Roche Diagnostics; support with conducting specificity and cross-reactivity experiments); and Stephen Weber (Roche Diagnostics; critical input).

Third-party medical writing support, under the direction of the authors, was provided by Thomas Burton (contract medical writer at Gardiner–Caldwell Communications, Macclesfield, UK) and was funded by Roche Diagnostics International Ltd (Rotkreuz, Switzerland).

COBAS, COBAS E and ELECSYS are trademarks of Roche.

